# Distinct Type 2 Diabetes Components Exert Contrasting Effects on Abdominal Aortic Aneurysm Development

**DOI:** 10.64898/2026.09.18.26363427

**Authors:** David DeVaro, Sridharan Raghavan, Venexia Walker, Tom Gaunt, Susanna Larsson, Michael G. Levin, Yae Hyun Rhee, Philip S. Tsao, Shuai Yuan, Scott M. Damrauer

## Abstract

**Background:** Type 2 diabetes (T2D) has been associated with reduced abdominal aortic aneurysm (AAA) risk and slower growth in observational studies, but the mechanism for this paradoxical association remains unclear. We hypothesized that T2D’s mechanistic heterogeneity obscures opposing, pathway-specific effects on AAA.

**Methods:** We performed two-sample Mendelian Randomization (MR) to investigate the effect of T2D (T2DGGI, 242,283 individuals with T2D and 1,569,730 without) on AAA (AAAgen, 37,214 individuals with AAA and 997,456 without) using 449 genetic instruments. We also performed MR within previously defined clusters representing different mechanisms of T2D. We extended the analysis to individual hyperglycemic, insulin-resistance, and insulin-sensitivity traits. Our hypothesis was tested in a porcine pancreatic elastase murine model of abdominal aortic aneurysm formation.

**Results:** Overall T2D liability showed no net association with AAA liability (OR 1.00, 95% CI 0.96-1.04, *P* = 0.90), albeit with marked instrument heterogeneity (Cochran’s Q = 1356, *P* = 5 x 10^-92^) and inconsistency across sensitivity analyses, with a notably protective MR-Egger result (OR 0.87, 95% CI 0.80-0.94, *P* = 0.0011) and non-zero intercept (*P* = 2 x 10^-4^). MR of T2D on AAA within mechanistic clusters showed opposing effects: lipodystrophy (OR 1.47, 95% CI 1.26-1.72) and obesity (OR 1.27, 95% CI 1.18-1.36) cluster membership associated with increased AAA risk, while beta-cell negative proinsulin (OR 0.83, 95% CI 0.76-0.90) and beta-cell positive proinsulin (OR 0.86, 95% CI 0.80-0.93) clusters were protective. This pattern was robust to clustering method. Traits indexing hyperglycemia (fasting glucose, 2-hour glucose, hemoglobin A1c) were associated with reduced AAA risk, while traits indexing insulin resistance (fasting insulin, BMI, waist-hip ratio, triglyceride-to-HDL ratio) were associated with increased AAA risk. In mice, those treated with a high fructose diet as a model for insulin resistance showed increased growth of the abdominal aorta compared with controls, whereas those treated with pancreatoxin streptozotocin as a model for isolated hyperglycemia showed blunted growth.

**Conclusion:** Distinct components of T2D physiology may have differential effects on AAA development. Whereas insulin resistance may promote AAA, hyperglycemia appears to protect against it. Our findings support insulin sensitization as a rational direction for AAA pharmacotherapy and suggest that the pathways mediating the protective effect of hyperglycemia may harbor additional therapeutic targets.

## Introduction

Abdominal aortic aneurysm (AAA) is a progressive dilation of the infrarenal aorta that remains clinically silent until rupture, a catastrophic event carrying a mortality of approximately 80%.^1^ Aortic aneurysm accounted for roughly 167,000 deaths worldwide in 2023,^2^ yet despite decades of investigation no pharmacological therapy has been shown to slow aneurysm growth or prevent rupture. More than a dozen placebo-controlled randomized trials of antibiotics, blood pressure lowering agents, antiplatelet drugs, mast-cell stabilizers, and fibrates have failed to demonstrate a convincing effect on aneurysm expansion.^3,4^ Management therefore remains limited to imaging surveillance until the aortic diameter reaches a threshold at which the risk of rupture exceeds the risk of operative repair, at which point open or endovascular repair is undertaken.^5^ This absence of medical therapy constitutes a major unmet clinical need and motivates the search for modifiable biological pathways governing aneurysm development.

One of the most striking observations in AAA epidemiology is its inverse association with type 2 diabetes (T2D). In a meta-analysis of sixteen prospective studies comprising over 16,000 incident cases among 4.5 million participants, diabetes was associated with a 42% lower risk of AAA.^6^ Individuals with diabetes also develop aneurysms that grow more slowly and are less prone to rupture.^7^ This association is notable because it runs opposite to the effect of diabetes on other forms of atherosclerotic cardiovascular disease; T2D confers roughly a two-fold excess risk across a wide range of vascular diseases, including coronary heart disease and stroke, and is a major risk factor for peripheral artery disease.^8^ The mechanism behind this divergent effect remains unclear, and further investigation could reveal targets for AAA pharmacotherapy.

Mendelian randomization (MR), which uses germline genetic variants as instruments for an exposure, offers a means to test whether the inverse association of T2D with AAA risk reflects a potentially causal effect. To date, however, genetic studies have not fully reconciled with the observational record. Two-sample MR analyses of genetic liability to T2D and glycemic traits have often reported null effects on AAA risk,^9–11^ even as MR studies of the thoracic aorta have found T2D and glycemic traits to be protective.^12,13^ One feature common to prior genetic analyses on AAA is that T2D was treated as a single, monolithic exposure. However, T2D is mechanistically heterogeneous, arising from distinct combinations of beta-cell dysfunction, insulin resistance, and adiposity, and recent large-scale efforts have resolved its genetic architecture into mechanistically defined clusters.^14,15^ A genetic instrument that averages across these mechanisms may obscure opposing effects on the aortic wall, yielding a null result even when constituent pathways exert real and divergent effects.

Here we revisited the association between T2D and AAA using the largest available genetic summary statistics. We first apply two-sample MR to explore the association between genetic liability to T2D and genetic liability to AAA, then partition that liability into mechanistically defined genetic clusters to test whether distinct biological components of T2D exert heterogeneous effects on AAA risk. We extend this analysis to individual glycemic traits and validate the resulting mechanistic hypothesis in a murine model of aneurysm formation (Figure 1).

**Figure 1.**
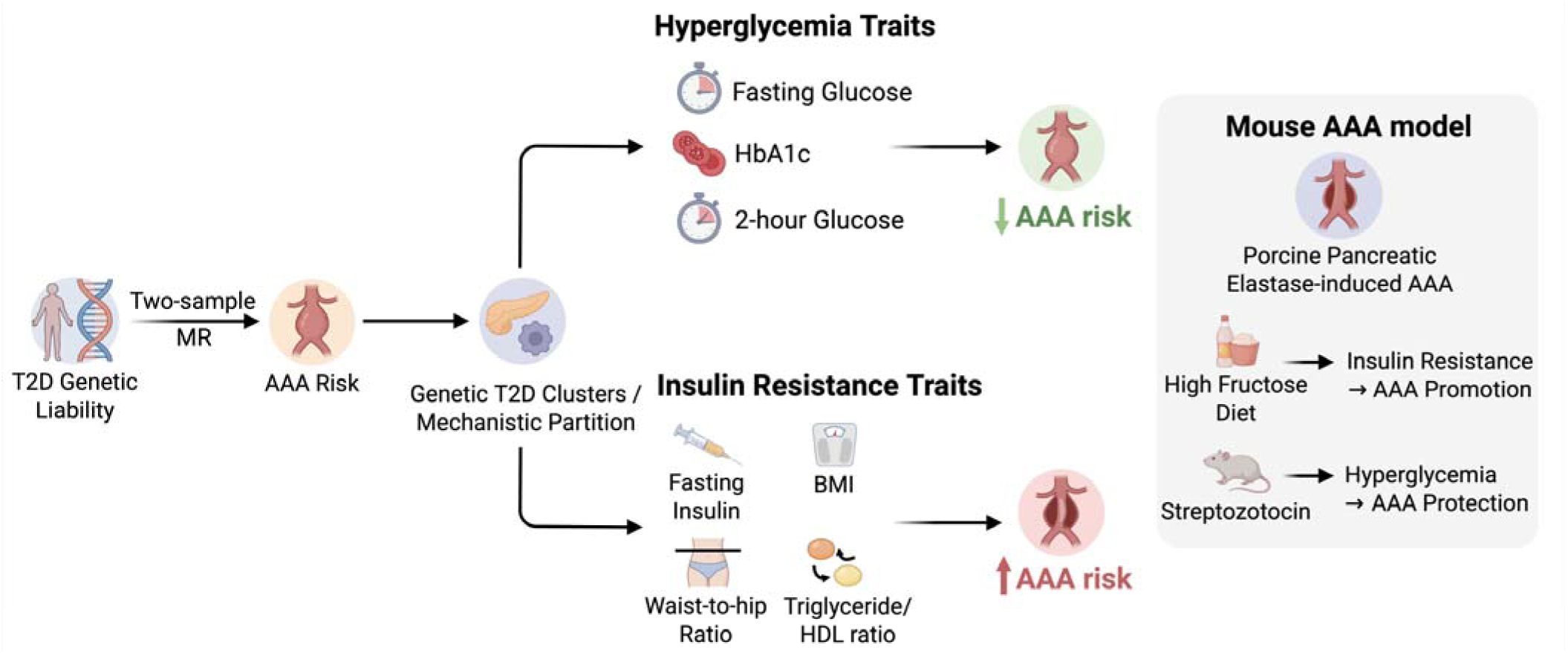
Overview of experiments. Two-sample mendelian randomization (MR) was first performed between type 2 diabetes (T2D) and abdominal aortic aneurysm (AAA). Two-sample MR was then performed within establish mechanistic clusters for T2D. In combination with two-sample MR between glycemic traits and AAA, this suggested that genetic liability to hyperglycemia protected against AAA whereas genetic liability to insulin resistance promoted AAA. This was also demonstrated in a porcine pancreatic elastase mouse model of AAA. Created with BioRender, https://BioRender.com/1lnal8h.

## Methods

### Statistical Genetics

#### Two-sample Mendelian randomization

We used two-sample Mendelian randomization to estimate the effects of type 2 diabetes (T2D) and glycemic traits on AAA using publicly available genome-wide association study (GWAS) summary statistics. We selected cohorts with the largest available sample size and used European-ancestry-specific summary statistics. Full details of the summary statistics, including sample sizes, ancestries, and accession identifiers, are provided in Supplementary Table 1.

Genetic liability to T2D was instrumented using summary statistics from the Type 2 Diabetes Global Genomics Initiative (T2DGGI) European-ancestry meta-analysis (242,283 cases and 1,569,730 controls).^14^ Effects on AAA were obtained from the AAAgen European-ancestry meta-analysis (37,214 cases and 997,456 controls).^16^ Glycemic traits were classified into hyperglycemic, insulin resistance, and insulin sensitivity groups. Hyperglycemic traits comprised fasting glucose adjusted for BMI (FGadjBMI), 2-hour glucose adjusted for BMI (2hGadjBMI),^17^ and glycated hemoglobin (HbA1c).^18^ Insulin-resistance traits comprised fasting insulin adjusted for BMI (FIadjBMI),^17^ body mass index (BMI), waist-to-hip ratio (WHR),^19^ and triglyceride-to-HDL-cholesterol ratio (TG:HDL).^20^ Insulin sensitivity was indexed by the modified Stumvoll insulin sensitivity index adjusted for BMI (ISIadjBMI).^21^

Instrumental variables were selected as variants with a minor allele frequency >0.01 reaching genome-wide significance (*P* < 5 x 10^-8^) with first stage F-statistic > 10. Linkage-disequilibrium (LD) clumping (r^2^ < 0.001) was performed using PLINK 1.9^22^ with a 1000 Genomes European reference panel.^23^ Exposure and outcome summary statistics were harmonized to align effect alleles prior to analysis. For T2D, this procedure yielded 449 instruments (Supplementary Table 2).

The inverse-variance weighted (IVW) method with multiplicative random effects was used as the primary analysis.^24^ To assess robustness to violations of MR assumptions, we additionally applied MR-Egger,^25^ weighted median,^26^ simple mode, weighted mode,^27^ contamination mixture,^28^ MR-PRESSO,^29^ and MR-RAPS^30^ methods, which rely on differing assumptions regarding instrument validity, and interpreted these estimators jointly as a panel rather than privileging any single sensitivity estimate (Supplementary Table 3). IVW, MR-Egger, weighted median, and weighted mode analyses were conducted using the TwoSampleMR package (version 0.6.25)^31^ in R (version 4.4.1). The contamination mixture method was implemented using the MendelianRandomization package (version 0.10.0).^32^ MR-PRESSO and MR-RAPS were run using the rondolab/MR-PRESSO and qingyuanzhao/mr.raps packages, respectively. Heterogeneity across instruments was quantified using Cochran’s Q statistic and is reported for completeness.

#### Mechanistic cluster analysis

To investigate heterogeneity in the effect of T2D-associated variants on AAA, we performed MR using two independently derived sets of mechanistic clusters of T2D-associated variants: hard clusters reported alongside the T2DGGI T2D GWAS in the main analysis^14^ and soft clusters reported by Kim et al in the supporting analysis.^15^ In either approach, genetic variants are assigned to clusters based on associations with an assortment of cardiometabolic traits, such as fasting glucose, systolic blood pressure, triglycerides, and LDL cholesterol. Hard and soft clustering differ in how variants are assigned. In hard clustering, each variant is assigned to a single cluster (here via k-means clustering across normalized cardiometabolic traits), whereas in soft clustering, each variant is assigned continuous weights across multiple clusters (here via Bayesian non-negative matrix factorization).

For the hard clusters, MR was performed separately on the subset of index variants assigned to each of the eight identified clusters. Cluster membership was taken as described and retained without re-selection based on association strength in the European-ancestry summary statistics. Variants with minor allele frequency < 0.01 were excluded. LD clumping (r^2^ < 0.001) was performed within each cluster rather than across the full variant set, to avoid removing cluster-specific instruments due to LD with variants assigned to other clusters (Supplementary Table 4).

For the soft clusters, each genome-wide significant T2D variant carries a continuous weight w for each of the ten identified clusters. LD clumping (r^2^ < 0.001) was performed once across the full variant set (Supplementary Table 5). Cluster-specific effect estimates were derived by scaling the inverse variance weights of the IVW regression by the cluster weights, such that variants with larger weights are upweighted. The soft-cluster results are interpreted directionally as a robustness check on the hard-cluster findings rather than as magnitude-matched estimates.

### Murine model

#### Mice

All animal protocols were approved by the Administrative Panel on Laboratory Animal Care at Stanford University (http://labanimals.stanford.edu/) and the VA Palo Alto Health Care System Institutional Animal Care and Use Committee and followed the National Institutes of Health and U.S. Department of Agriculture Guidelines for Care and Use of Animals in Research. Eight-week old C57BL/6J were purchased from The Jackson Laboratory (Bar Harbor, ME, USA) and maintained in a temperature-controlled room (22 °C ± 1 °C) with a 12-hour light/dark cycle and *ad libitum* access to water. Control animals were fed normal Purina chow (No. 5012). A subset of animals was fed high-fructose chow (60% fructose, 11% fat, and 22% protein; Teklad Labs) for two weeks prior to PPE surgery and continued throughout the 4-week follow up period. The fructose-enriched diet leads to insulin resistance, hyperinsulinemia, and hypertriglyceridemia.^33,34^ To induce hyperglycemic state, a third group of animals were administered five daily intraperitoneal injections of the islet toxin, streptozotocin (STZ), 50 mg/kg in citrate buffer (Sigma Aldrich) starting two weeks before PPE surgery.^35^ Hyperglycemia was defined by casual blood glucose levels of ≥300 mg/dL before AAA creation and at sacrifice.

#### Porcine pancreatic elastase (PPE) infusion model

The PPE infusion model to induce mouse AAA was performed as previously described at 10 weeks of age.^36^ The proximal and distal aorta were temporarily ligated or clamped, followed by an aortotomy above the iliac bifurcation. A catheter was used to infuse the aorta for 5 minutes at 120 mmHg with saline, or saline containing type I porcine pancreatic elastase (2.5 U/mL; Sigma Aldrich), and the aortotomy was then repaired. The procedure results in a progressive aneurysm between the left renal artery and the bifurcation that is followed for 28 days post-surgery.

#### AAA growth and blood pressure monitoring

AAA development in both models was monitored via B-mode ultrasound (Vevo 2100® High-Resolution In Vivo Micro-Imaging System; VisualSonics, Toronto, Canada) at baseline and day 3, 7, 14, 21, and 28 post-surgery. Mice were studied under isoflurane anesthesia and aortic diameters were assessed at maximum diameter in the systolic cardiac phase.

## Results

### Null albeit heterogeneous association between genetic liability to type 2 diabetes and AAA

Using 449 genetic IVs, two-sample MR testing the association of overall genetic liability for T2D to genetic liability for AAA yielded no association using the inverse-variance weighted (IVW) method (OR of AAA 1.00 per unit higher log odds of diabetes, 95% CI 0.96-1.04, *P* = 0.90). However, this null estimate masked pronounced heterogeneity across instruments (Cochran’s Q = 1356, *P* = 5 x 10^-92^). Notably, several pleiotropy-robust estimators diverged markedly from the IVW result, including the MR-Egger result (OR 0.87, 95% CI 0.80-0.94, *P* = 0.0011), with a non-zero MR-Egger intercept (*P* = 2 x 10^-4^) (Figure 2, Supplementary Table 6).

**Figure 2.**
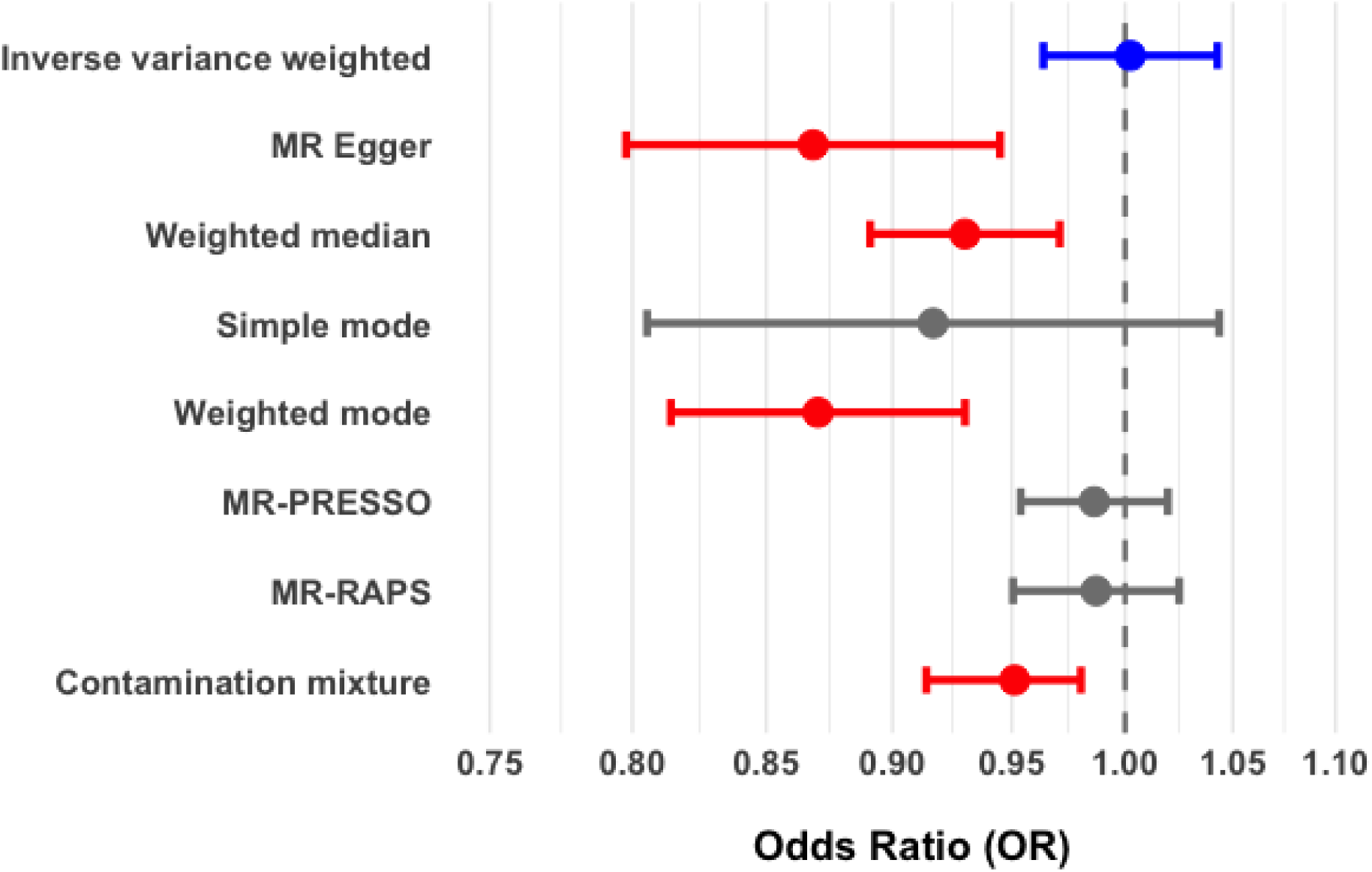
Two-sample mendelian randomization experiment for the effect of T2D on AAA. Effect estimates are reported across all utilized MR methods with 95% confidence intervals, with odds ratios reported for AAA per unit log odds T2D. The inverse variance weighted estimate is included in blue, as the primary analysis. For the remaining sensitivity analyses, statistically significant results are reported in red and nonsignificant results reported in gray.

To further quantify the relationship between more specific T2D mechanisms and AAA, we then performed MR using genetic instruments for each of the 8 T2DGGI hard clusters, which segregated into opposing effects on AAA (odds ratios for AAA per unit log odds T2D). Genetic liability for T2D for variants in the lipodystrophy cluster (OR 1.47, 95% CI 1.26-1.72, *P* = 1.1 x 10^-6^) and the obesity cluster (OR 1.27, 95% CI 1.18-1.36, *P* = 2.0 x 10^-10^) was associated with increased genetic liablity for AAA. In contrast, both beta-cell clusters were protective: beta cell - PI (OR 0.83, 95% CI 0.76-0.90, *P* = 2.7 x 10^-5^) and beta cell +PI (OR 0.86, 95% CI 0.80-0.93, *P* = 2.7 x 10^-4^). The remaining clusters showed weaker or null associations: metabolic syndrome (OR 0.90, 95% CI 0.82-0.99, *P* = 0.038), residual glycaemic (OR 0.99, 95% CI 0.92-1.07, *P* = 0.82), body fat (OR 1.05, 95% CI 0.98-1.12, *P* = 0.21), and liver/lipid metabolism (OR 0.30, 95% CI 0.09-1.04, *P* = 0.058; instrumented by only three variants) (Figure 3, Supplementary Table 7, Supplementary Figure 1). The directionally opposing effects of the lipodystrophy/obesity and beta-cell clusters were consistent across pleiotropy-robust estimators (Supplementary Figure 2), indicating that, unlike for the pooled estimate, these within-cluster IVW estimates were not substantially affected by residual heterogeneity.

**Figure 3.**
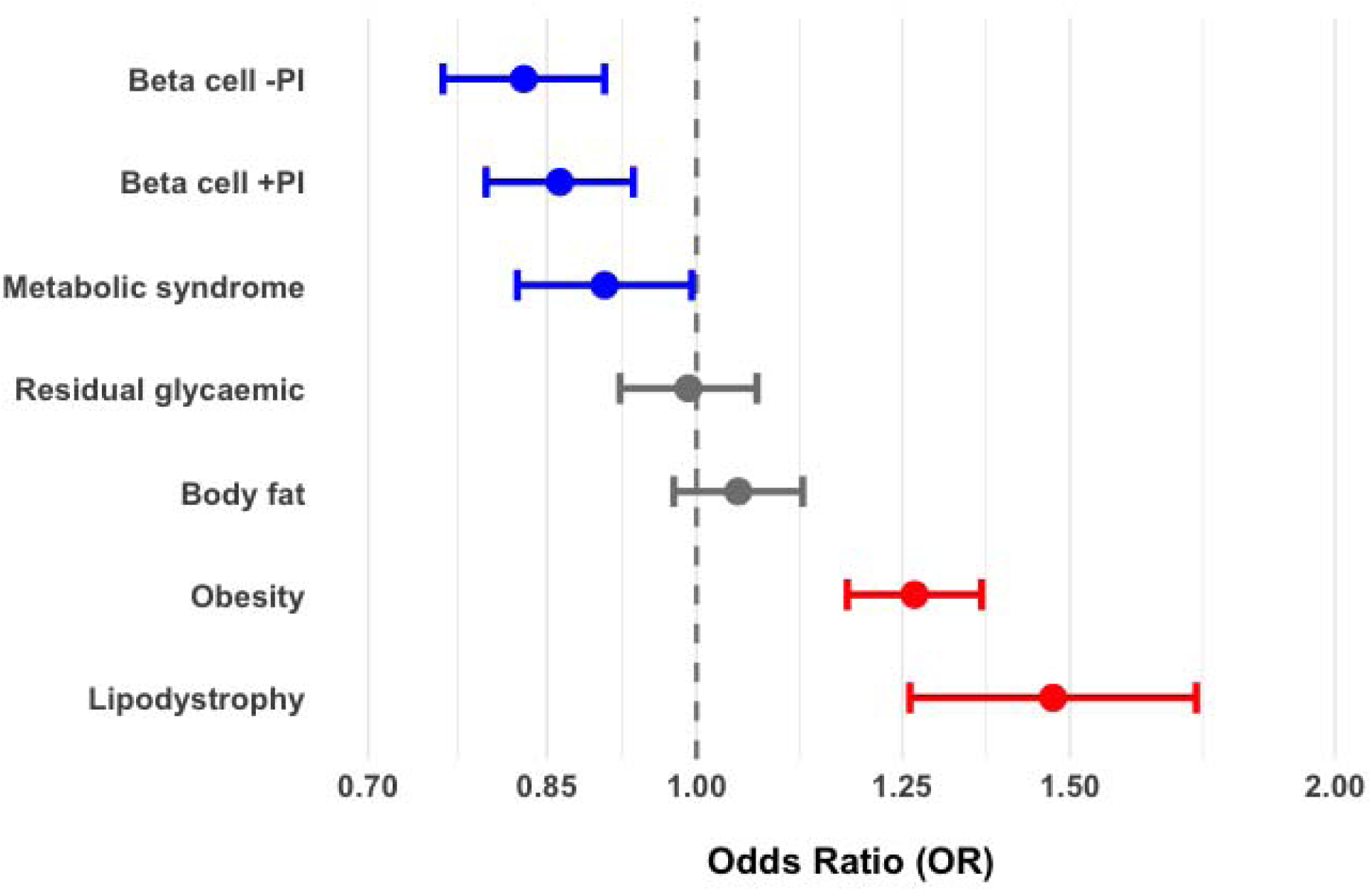
Inverse-variance weighted effect estimates of Suzuki et al. T2D hard clusters on AAA risk. IVW effect estimates for the effect of T2D on AAA are reported for each cluster with 95% confidence intervals, with odds ratios reported for AAA per unit log odds T2D. Clusters associated with decreased AAA risk are depicted in blue whereas those associated with increased AAA risk are depicted in red. Nonsignificant results reported in gray. The Liver/lipid metabolism cluster only comprised 3 SNPs and is not included here.

We observed directionally concordant results using independently derived soft clusters. Beta-cell and proinsulin clusters were protective--Beta cell 1 (OR 0.90, 95% CI 0.87-0.93, *P* = 6.8 x 10^-9^), Beta cell 2 (OR 0.94, 95% CI 0.89-0.98, *P* = 4.3 x 10^-3^), and Proinsulin (OR 0.87, 95% CI 0.82-0.93, *P* = 2.7 x 10^-5^)--whereas the obesity (OR 1.14, 95% CI 1.07-1.22, *P* = 5.6 x 10^-5^), lipodystrophy (OR 1.24, 95% CI 1.18-1.29, *P* = 1.9 x 10^-21^), and hyper-insulin-secretion (OR 1.13, 95% CI 1.05-1.21, *P* = 7.3 x 10^-4^) clusters associated with increased liability for AAA. The remaining soft clusters (lipoprotein A, liver/lipid, ALP negative, SHBG) showed no significant association (Supplementary Table 8, Supplementary Figure 3).

### An insulin-resistance versus hyperglycemia axis predicts trait-level effects on AAA

We next investigated the effects of different glycemic traits on AAA. Genetic liability to traits indexing hyperglycemia was protective: fasting glucose adjusted for BMI (FGadjBMI, OR 0.78, 95% CI 0.65-0.95, *P* = 0.012) and 2-hour glucose adjusted for BMI (2hGadjBMI, OR 0.81, 95% CI 0.68-0.95, *P* = 0.009) were each significantly associated with reduced liability for AAA under IVW, and glycated haemoglobin (HbA1c) was concordantly protective across pleiotropy-robust estimators though null under IVW (OR 0.97, 95% CI 0.92-1.02, *P* = 0.25). Conversely, genetic liability for traits indexing insulin resistance was strongly associated with increased liability for AAA: fasting insulin adjusted for BMI (FIadjBMI, OR 2.06, 95% CI 1.33-3.17, *P* = 0.0011), body mass index (OR 1.48, 95% CI 1.38-1.60, *P* = 1.9 x 10^-26^), waist-hip ratio (OR 1.28, 95% CI 1.13-1.46, *P* = 2 x 10^-4^), and triglyceride-to-HDL ratio (OR 1.63, 95% CI 1.48-1.79, *P* = 8 x 10^-24^). Consistent with this axis, genetic liability to higher insulin sensitivity (ISIadjBMI), the inverse of insulin resistance, was protective (OR 0.67, 95% CI 0.49-0.92, *P* = 0.014) (Figure 4, Supplementary Table 6, Supplementary Figure 4).

**Figure 4.**
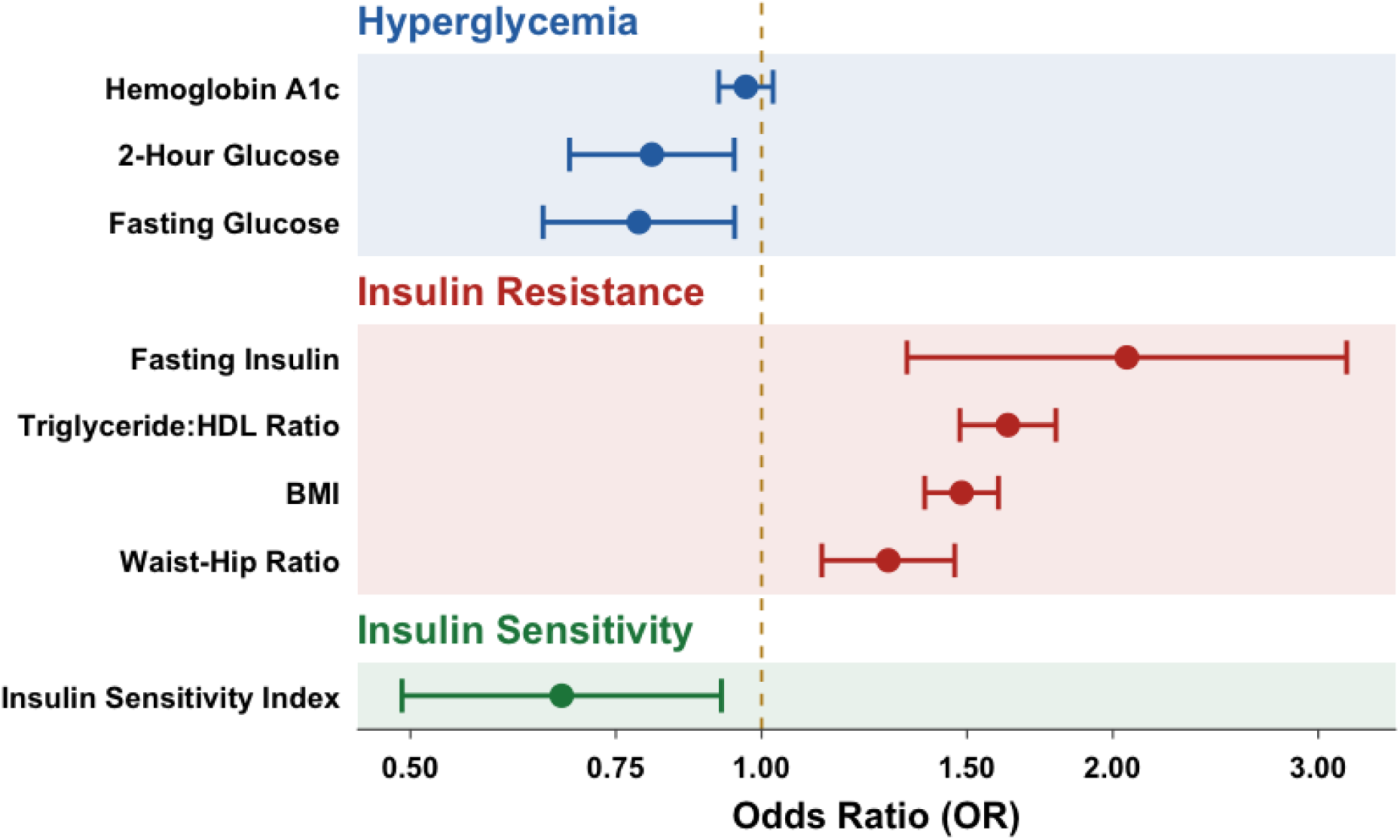
Inverse-variance weighted effect estimates of glycemic traits on AAA. IVW estimates for the effects of an assortment of glycemic traits on AAA are reported with 95% confidence intervals. Traits are divided into those indexing hyperglycemia (Hemoglobin A1c, 2-hour glucose adjusted for BMI, and fasting glucose adjusted for BMI), insulin resistance (fasting insulin adjusted for BMI, triglyceride-to-HDL ratio, BMI, waist-to-hip ratio), and insulin sensitivity (modified Stumvoll insulin sensitivity index). Effect estimates are in different units and should be interpreted according to directional effect rather than through comparison of magnitudes.

### Murine validation

Using an established model of AAA induction, the porcine-pancreatic-elastase (PPE) infusion model in 10 week old male C57Bl/6J mice, we investigated whether two components of T2D, insulin resistance/hyperinsulinemia and hyperglycemia, had differing effects on the development of experimental AAA. B-mode ultrasound imaging performed 3, 7, 14, 21 and 28 days after PPE infusion showed a significant increase in expansion of the abdominal aortic diameter (AAD) from day 21 until day 28 in fructose-fed compared to normal chow-fed mice. By contrast, streptozotocin-induced hyperglyemia was associated with a significant reduction in AAD starting at day 7 and persisted throughout the followup period (Figure 5, Supplementary Table 9).

**Figure 5.**
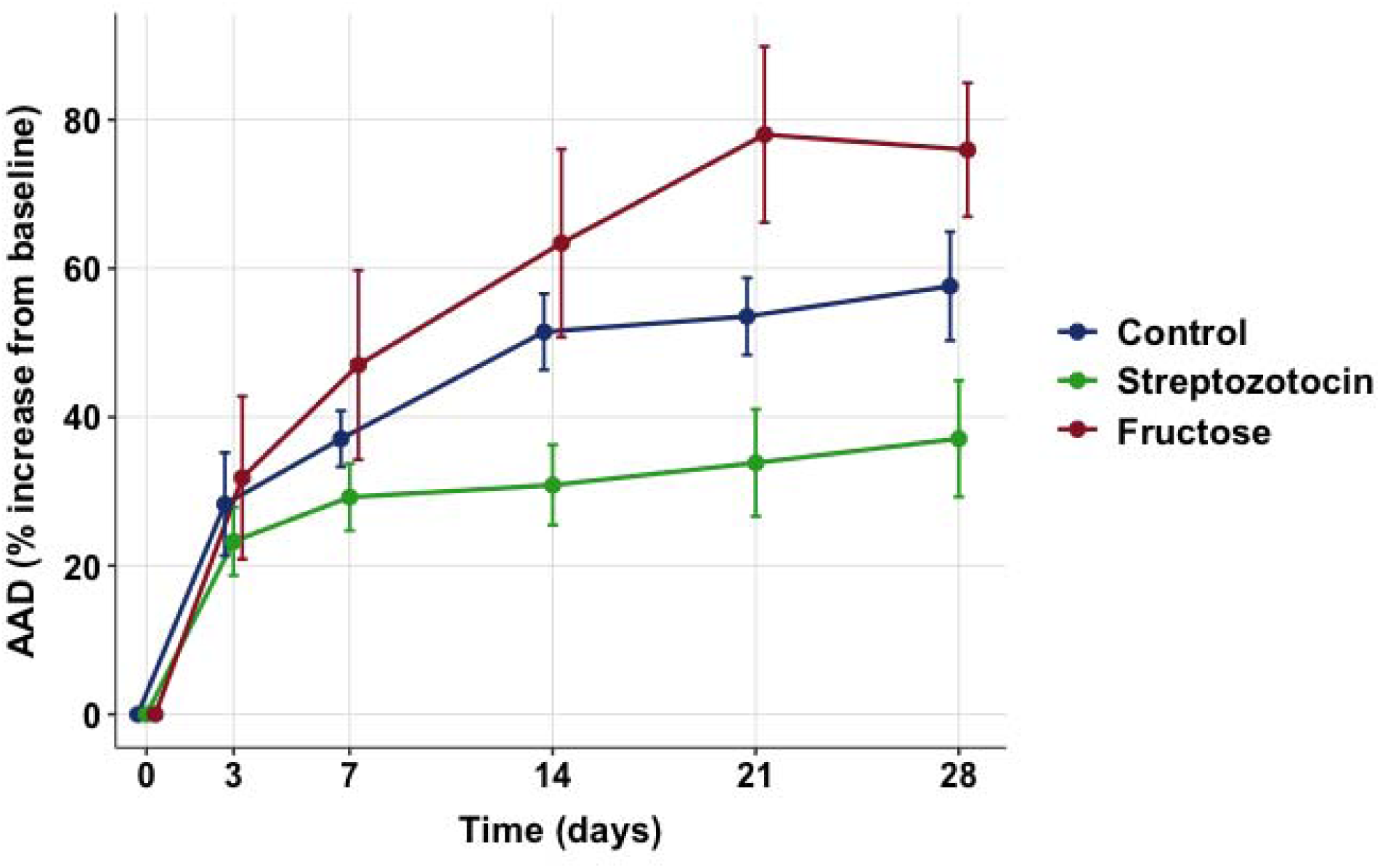
Increase in abdominal aortic diameter in mice treated with porcine pancreatic elastase. Average percent increase from baseline abdominal aortic diameter (AAD) after treatment with porcine pancreatic elastase is reported at days 3, 7, 14, 21, and 28 for treatment with control diet (blue), high fructose diet (red), and streptozotocin (green). Error bars are included as 95% confidence intervals.

## Discussion

In this study, we found that genetic liability to T2D exerts no detectable net effect on AAA risk under a conventional pooled analysis, but that this null estimate conceals substantial, mechanistically structured heterogeneity. Partitioning T2D liability into established mechanistic clusters resolved this heterogeneity into opposing effects: clusters reflecting insulin resistance-- lipodystrophy and obesity--promoted AAA, whereas clusters reflecting beta-cell dysfunction and impaired insulin secretion were protective. This pattern was reproduced using an independently derived soft-clustering approach and predicted a physiological dichotomy that we confirmed using genetic instruments for individual glycemic traits: hyperglycemia-indexing traits were protective, while insulin-resistance-indexing traits promoted AAA. Mouse models of AAA further supported this dichotomy.

Observational studies have long and reproducibly associated diabetes with reduced AAA prevalence, growth, and rupture, yet prior two-sample MR studies using genetic liability to T2D often found no protective effect.^9,11^ Our results offer a resolution: the net effect of T2D liability on liability to AAA is the sum of opposing, mechanism-specific effects, such that averaging across these mechanisms yields a null or unstable pooled estimate. Notably, several of our pleiotropy-robust estimators for overall T2D were directionally protective, consistent with the possibility that the net effect of T2D liability is indeed protective on balance. Additionally, we suggest that the net effect decomposes into a harmful insulin-resistance component and a protective hyperglycemia component.

The interpretation we advance rests on the established physiology of T2D. Insulin resistance in peripheral tissues and progressive beta-cell failure are the two core, partially separable pathophysiologic defects of T2D,^37,38^ and the genetic clusters we employed were constructed precisely to distinguish these mechanisms. Mapping our results onto this framework, the clusters that promote AAA (lipodystrophy, obesity, and--in the soft-cluster analysis--hyper-insulin secretion) share a physiology of insulin resistance, while the protective clusters (beta-cell ±PI, proinsulin) reflect impaired insulin secretion and consequent relative hyperglycemia with relatively lower circulating insulin. That genetic instruments for individual traits--constructed entirely independently of the cluster analysis--reproduce the same split provides convergent support. Fasting and post-load glucose were protective, multiple insulin-resistance indices (fasting insulin, BMI, waist-hip ratio, triglyceride-to-HDL ratio) were harmful, and higher insulin sensitivity was protective.

This dichotomy is consistent with a body of prior work that, when viewed individually, has appeared contradictory. On the hyperglycemia side, clinical studies have found that higher glucose exposure tracks with less aneurysm disease: fasting glucose concentration is inversely associated with infra-renal aortic diameter,^39^ and glycated hemoglobin is inversely associated with AAA growth rate.^40^ On the insulin-resistance side, the limited available clinical data point the opposite way: markers of insulin resistance and endogenous insulin secretion--C-peptide, insulin, and the HOMA2-derived insulin resistance index--are significantly higher in patients with larger than with smaller aneurysms.^41^ Our findings provide support for both arms of this fragmented picture.

The mechanisms underlying these opposing effects remain incompletely understood and represent a rich avenue for further investigation. For the protective hyperglycemia effect, one candidate is the formation of advanced glycation end-products (AGEs) and consequent non-enzymatic collagen cross-linking, which may stabilize the aortic extracellular matrix against proteolytic degradation.^7^ Interestingly, pharmacologic inhibition of AGE formation or AGE-matrix cross-linking reverses the protection conferred by hyperglycemia in diabetic mice without altering serum glucose, directly implicating matrix stabilization in the protective phenotype.^42^ Additional glucose-dependent mechanisms, including modulation of fibrinolysis, inflammation, and mural angiogenesis, have also been proposed.^7,43^

For the harmful insulin-resistance effect, the dyslipidemia intrinsic to insulin-resistant states is a plausible partial mediator. The lipodystrophy cluster, our strongest harmful genetic signal, is defined by elevated triglycerides, reduced HDL cholesterol, and elevated waist-hip ratio, and our triglyceride-to-HDL instrument was among the most strongly AAA-promoting traits tested. This aligns with robust genetic evidence that circulating lipids and lipid-lowering drug targets are causally associated with AAA,^44^ including our prior meta-analysis identifying multiple lipid loci and PCSK9 as a therapeutic target.^16^ The harmful effect of insulin resistance on AAA may therefore be mediated, at least in part, through its characteristic atherogenic lipid profile.

These findings have translational resonance. There is currently no approved pharmacotherapy to limit AAA growth, and numerous trials of blood-pressure-lowering, antibiotic, antiplatelet, and lipid-lowering agents have not convincingly altered disease progression.^3,4^ Our data, by implicating insulin resistance as harmful, support the rationale for insulin-sensitizing strategies. Metformin, in particular, has been associated with slower AAA growth in multiple observational cohorts^45^ and is the subject of several ongoing randomized trials in non-diabetic patients with small AAA.^46–48^ Our genetic results provide independent support for this therapeutic hypothesis and, more broadly, suggest that the protective hyperglycemia axis may itself harbor exploitable targets. For example, although AGEs have been strongly implicated in diabetes-associated cardiovascular disease,^49^ strategies that locally recapitulate AGE-mediated matrix stabilization without the systemic harms of chronic hyperglycemia could offer a potential therapeutic avenue. Notably, matrix-stabilizing approaches are already advancing toward the clinic through a mechanistically distinct route: pentagalloyl glucose, a polyphenol that binds elastin and collagen and protects them from proteolytic degradation, suppresses experimental AAA in preclinical models^50,51^ and is now being evaluated as a local endovascular therapy for small-to-medium AAA with an ongoing randomized controlled trial.^52^

Several limitations warrant mention. Firstly, our analyses were restricted to individuals of European ancestry due to MR methodologic constraints and thus do not benefit from the consideration of diverse populations. Second, the hard cluster definitions were derived in part from multi-ancestry data while our instruments used European-ancestry summary statistics. However, directionality of estimates was confirmed with independent soft-cluster estimates, although we emphasize that the magnitude of these estimates is not interpretable due to the soft cluster weighting procedure required. Third, our murine models may not well reflect T2D physiology in humans, nor does the porcine pancreatic elastase model perfectly model AAA pathogenesis. Fourth, the mechanisms underlying the observed effects remain to be established, and the candidate mediators we discuss are hypotheses that cannot be further validated from our analysis. Finally, MR rests on a series of assumptions--instrument relevance, independence, and exclusion restriction--that cannot be fully verified; we mitigated these through the use of multiple pleiotropy-robust estimators interpreted as a panel.

## Conclusion

The heterogeneous relationship between T2D and AAA reflects an opposition between insulin resistance, which promotes aneurysm development, and hyperglycemia, which protects against it. This framework reconciles the apparent discordance in prior observational data, unifies a fragmentary experimental literature, and identifies insulin sensitization as a rational direction for the development of urgently needed AAA pharmacotherapy. Defining the mechanisms by which hyperglycemia protects against AAA may reveal additional therapeutic targets.

## Data Availability

All data produced in the present work are contained in the manuscript and supplementary data.

https://www.ebi.ac.uk/gwas/

http://doi.org/10.1038/s41586-024-07019-6

http://doi.org/10.1007/s00125-022-05848-6

http://doi.org/10.1038/s41588-023-01510-y

## References

1. Reimerink JJ, Van Der Laan MJ, Koelemay MJ, Balm R, Legemate DA. Systematic review and meta-analysis of population-based mortality from ruptured abdominal aortic aneurysm. Br J Surg. 2013;100(11):1405–1413. doi:10.1002/bjs.9235

2. Stark BA, DeCleene NK, Desai EC, Hsu JM, Johnson CO, Lara-Castor L, et al. Global, Regional, and National Burden of Cardiovascular Diseases and Risk Factors in 204 Countries and Territories, 1990-2023. JACC. 2025;86(22):2167–2243. doi:10.1016/j.jacc.2025.08.015

3. Golledge J, Thanigaimani S, Powell JT, Tsao PS. Pathogenesis and management of abdominal aortic aneurysm. Eur Heart J. 2023;44(29):2682–2697. doi:10.1093/eurheartj/ehad386

4. Golledge J, Moxon JV, Singh TP, Bown MJ, Mani K, Wanhainen A. Lack of an effective drug therapy for abdominal aortic aneurysm. J Intern Med. 2020;288(1):6–22. doi:10.1111/joim.12958

5. Wanhainen A, Verzini F, Van Herzeele I, Allaire E, Bown M, Cohnert T, et al. Editor’s Choice – European Society for Vascular Surgery (ESVS) 2019 Clinical Practice Guidelines on the Management of Abdominal Aorto-iliac Artery Aneurysms. Eur J Vasc Endovasc Surg. 2019;57(1):8–93. doi:10.1016/j.ejvs.2018.09.020

6. Aune D, Schlesinger S, Norat T, Riboli E. Diabetes mellitus and the risk of abdominal aortic aneurysm: A systematic review and meta-analysis of prospective studies. J Diabetes Complications. 2018;32(12):1169–1174. doi:10.1016/j.jdiacomp.2018.09.009

7. Patel K, Zafar MA, Ziganshin BA, Elefteriades JA. Diabetes Mellitus: Is It Protective against Aneurysm? A Narrative Review. Cardiology. 2018;141(2):107–122. doi:10.1159/000490373

8. The Emerging Risk Factors Collaboration. Diabetes mellitus, fasting blood glucose concentration, and risk of vascular disease: a collaborative meta-analysis of 102 prospective studies. The Lancet. 2010;375(9733):2215–2222. doi:10.1016/S0140-6736(10)60484-9

9. Morris DR, Jones GT, Holmes MV, Bown MJ, Bulbulia R, Singh TP, et al. Genetic Predisposition to Diabetes and Abdominal Aortic Aneurysm: A Two Stage Mendelian Randomisation Study. Eur J Vasc Endovasc Surg. 2022;63(3):512–519. doi:10.1016/j.ejvs.2021.10.038

10. Niu Z, Cao L, Guo W, Zhang H. Associations between Type 2 Diabetes Mellitus, Metabolic Traits, and Abdominal Aortic Aneurysm: A Cross-Ethnic Mendelian Randomization Analysis. Ann Vasc Surg. 2025;110:405–413. doi:10.1016/j.avsg.2024.07.105

11. Van’T Hof FN, Vaucher J, Holmes MV, De Wilde A, Baas AF, Blankensteijn JD, et al. Genetic variants associated with type 2 diabetes and adiposity and risk of intracranial and abdominal aortic aneurysms. Eur J Hum Genet. 2017;25(6):758–762. doi:10.1038/ejhg.2017.48

12. Zhang Y, Li Y, Dai X, Lin H, Ma L. Type 2 diabetes has a protective causal association with thoracic aortic aneurysm: a Mendelian randomization study. Diabetol Metab Syndr. 2023;15(1):120. doi:10.1186/s13098-023-01101-1

13. Daria T, Iyer K, Alkhairo H, Kho PF, Suzuki K, Hatzikotoulas K, et al. Mendelian Randomization Suggests a Causal Link Between Glycemic Traits and Thoracic Aortic Structures and Diseases. JACC Basic Transl Sci. 2025;10(11):101390. doi:10.1016/j.jacbts.2025.101390

14. Suzuki K, Hatzikotoulas K, Southam L, Taylor HJ, Yin X, Lorenz KM, et al. Genetic drivers of heterogeneity in type 2 diabetes pathophysiology. Nature. 2024;627(8003):347–357. doi:10.1038/s41586-024-07019-6

15. Kim H, Westerman KE, Smith K, Chiou J, Cole JB, Majarian T, et al. High-throughput genetic clustering of type 2 diabetes loci reveals heterogeneous mechanistic pathways of metabolic disease. Diabetologia. 2023;66(3):495–507. doi:10.1007/s00125-022-05848-6

16. Roychowdhury T, Klarin D, Levin MG, Spin JM, Rhee YH, Deng A, et al. Genome-wide association meta-analysis identifies risk loci for abdominal aortic aneurysm and highlights PCSK9 as a therapeutic target. Nat Genet. 2023;55(11):1831–1842. doi:10.1038/s41588-023-01510-y

17. Chen J, Spracklen CN, Marenne G, Varshney A, Corbin LJ, Luan J, et al. The trans-ancestral genomic architecture of glycemic traits. Nat Genet. 2021;53(6):840–860. doi:10.1038/s41588-021-00852-9

18. Karczewski KJ, Gupta R, Kanai M, Lu W, Tsuo K, Wang Y, et al. Pan-UK Biobank genome-wide association analyses enhance discovery and resolution of ancestry-enriched effects. Nat Genet. 2025;57(10):2408–2417. doi:10.1038/s41588-025-02335-7

19. Pulit SL, Stoneman C, Morris AP, Wood AR, Glastonbury CA, Tyrrell J, et al. Meta-analysis of genome-wide association studies for body fat distribution in 694 649 individuals of European ancestry. Hum Mol Genet. 2019;28(1):166–174. doi:10.1093/hmg/ddy327

20. DeForest N, Wang Y, Zhu Z, Dron JS, Koesterer R, Natarajan P, et al. Genome-wide discovery and integrative genomic characterization of insulin resistance loci using serum triglycerides to HDL-cholesterol ratio as a proxy. Nat Commun. 2024;15(1):8068. doi:10.1038/s41467-024-52105-y

21. Williamson A, Norris DM, Yin X, Broadaway KA, Moxley AH, Vadlamudi S, et al. Genome-wide association study and functional characterization identifies candidate genes for insulin-stimulated glucose uptake. Nat Genet. 2023;55(6):973–983. doi:10.1038/s41588-023-01408-9

22. Chang CC, Chow CC, Tellier LC, Vattikuti S, Purcell SM, Lee JJ. Second-generation PLINK: rising to the challenge of larger and richer datasets. Gigascience. 2015;4(1):s13742-015-0047-0048. doi:10.1186/s13742-015-0047-8

23. The 1000 Genomes Project Consortium, Corresponding authors, Auton A, Abecasis GR, Steering committee, Altshuler DM, et al. A global reference for human genetic variation. Nature. 2015;526(7571):68-74. doi:10.1038/nature15393

24. Burgess S, Butterworth A, Thompson SG. Mendelian Randomization Analysis With Multiple Genetic Variants Using Summarized Data. Genet Epidemiol. 2013;37(7):658–665. doi:10.1002/gepi.21758

25. Bowden J, Davey Smith G, Burgess S. Mendelian randomization with invalid instruments: effect estimation and bias detection through Egger regression. Int J Epidemiol. 2015;44(2):512–525. doi:10.1093/ije/dyv080

26. Bowden J, Davey Smith G, Haycock PC, Burgess S. Consistent Estimation in Mendelian Randomization with Some Invalid Instruments Using a Weighted Median Estimator. Genet Epidemiol. 2016;40(4):304–314. doi:10.1002/gepi.21965

27. Hartwig FP, Davey Smith G, Bowden J. Robust inference in summary data Mendelian randomization via the zero modal pleiotropy assumption. Int J Epidemiol. 2017;46(6):1985–1998. doi:10.1093/ije/dyx102

28. Burgess S, Foley CN, Allara E, Staley JR, Howson JMM. A robust and efficient method for Mendelian randomization with hundreds of genetic variants. Nat Commun. 2020;11(1):376. doi:10.1038/s41467-019-14156-4

29. Verbanck M, Chen CY, Neale B, Do R. Detection of widespread horizontal pleiotropy in causal relationships inferred from Mendelian randomization between complex traits and diseases. Nat Genet. 2018;50(5):693–698. doi:10.1038/s41588-018-0099-7

30. Zhao Q, Wang J, Hemani G, Bowden J, Small DS. Statistical inference in two-sample summary-data Mendelian randomization using robust adjusted profile score. Ann Stat. 2020;48(3). doi:10.1214/19-AOS1866

31. Hemani G, Zheng J, Elsworth B, Wade KH, Haberland V, Baird D, et al. The MR-Base platform supports systematic causal inference across the human phenome. eLife. 2018;7:e34408. doi:10.7554/eLife.34408

32. Yavorska OO, Burgess S. MendelianRandomization: an R package for performing Mendelian randomization analyses using summarized data. Int J Epidemiol. 2017;46(6):1734–1739. doi:10.1093/ije/dyx034

33. Tsao PS, Niebauer J, Buitrago R, Lin PS, Wang BY, Cooke JP, et al. Interaction of diabetes and hypertension on determinants of endothelial adhesiveness. Arterioscler Thromb Vasc Biol. 1998;18(6):947–953. doi:10.1161/01.atv.18.6.947

34. Montgomery MK, Fiveash CE, Braude JP, Osborne B, Brown SHJ, Mitchell TW, et al. Disparate metabolic response to fructose feeding between different mouse strains. Sci Rep. 2015;5:18474. doi:10.1038/srep18474

35. Terashima M, Ehara S, Yang E, Kosuge H, Tsao PS, Quertermous T, et al. In vivo bioluminescence imaging of inducible nitric oxide synthase gene expression in vascular inflammation. Mol Imaging Biol. 2011;13(6):1061–1066. doi:10.1007/s11307-010-0451-5

36. Azuma J, Asagami T, Dalman R, Tsao PS. Creation of murine experimental abdominal aortic aneurysms with elastase. J Vis Exp JoVE. 2009;(29):1280. doi:10.3791/1280

37. Kahn SE. The relative contributions of insulin resistance and beta-cell dysfunction to the pathophysiology of Type 2 diabetes. Diabetologia. 2003;46(1):3–19. doi:10.1007/s00125-002-1009-0

38. DeFronzo RA. From the Triumvirate to the Ominous Octet: A New Paradigm for the Treatment of Type 2 Diabetes Mellitus. Diabetes. 2009;58(4):773–795. doi:10.2337/db09-9028

39. Le MTQ, Jamrozik K, Davis TME, Norman PE. Negative Association between Infra-renal Aortic Diameter and Glycaemia: The Health In Men Study. Eur J Vasc Endovasc Surg. 2007;33(5):599–604. doi:10.1016/j.ejvs.2006.12.017

40. Kristensen KL, Dahl M, Rasmussen LM, Lindholt JS. Glycated Hemoglobin Is Associated With the Growth Rate of Abdominal Aortic Aneurysms: A Substudy From the VIVA (Viborg Vascular) Randomized Screening Trial. Arterioscler Thromb Vasc Biol. 2017;37(4):730–736. doi:10.1161/ATVBAHA.116.308874

41. Lareyre F, Moratal C, Zereg E, Carboni J, Panaïa-Ferrari P, Bayer P, et al. Association of abdominal aortic aneurysm diameter with insulin resistance index. Biochem Medica. 2018;28(3):030702. doi:10.11613/BM.2018.030702

42. Li Y, Zheng X, Guo J, Samura M, Ge Y, Zhao S, et al. Treatment With Small Molecule Inhibitors of Advanced Glycation End Products Formation and Advanced Glycation End Products Mediated Collagen Cross Linking Promotes Experimental Aortic Aneurysm Progression in Diabetic Mice. J Am Heart Assoc. 2023;12(10):e028081. doi:10.1161/JAHA.122.028081

43. Raffort J, Lareyre F, Clément M, Hassen-Khodja R, Chinetti G, Mallat Z. Diabetes and aortic aneurysm: current state of the art. Cardiovasc Res. 2018;114(13):1702–1713. doi:10.1093/cvr/cvy174

44. Harrison SC, Holmes MV, Burgess S, Asselbergs FW, Jones GT, Baas AF, et al. Genetic Association of Lipids and Lipid Drug Targets With Abdominal Aortic Aneurysm: A Meta-analysis. JAMA Cardiol. 2018;3(1):26. doi:10.1001/jamacardio.2017.4293

45. Thanigaimani S, Singh TP, Unosson J, Phie J, Moxon J, Wanhainen A, et al. Editor’s Choice – Association Between Metformin Prescription and Abdominal Aortic Aneurysm Growth and Clinical Events: a Systematic Review and Meta-Analysis. Eur J Vasc Endovasc Surg. 2021;62(5):747–756. doi:10.1016/j.ejvs.2021.06.013

46. Wanhainen A, Unosson J, Mani K, Gottsäter A, Olsson KW, Björck M, et al. The Metformin for Abdominal Aortic Aneurysm Growth Inhibition (MAAAGI) Trial. Eur J Vasc Endovasc Surg. 2021;61(4):710–711. doi:10.1016/j.ejvs.2020.11.048

47. Dalman RL, Lu Y, Mahaffey KW, Chase AJ, Stern JR, Chang RW. Background and Proposed Design for a Metformin Abdominal Aortic Aneurysm Suppression Trial. Vasc Endovasc Rev. 2020;3:e08. doi:10.15420/ver.2020.03

48. Golledge J, Arnott C, Moxon J, Monaghan H, Norman R, Morris D, et al. Protocol for the Metformin Aneurysm Trial (MAT): a placebo-controlled randomised trial testing whether metformin reduces the risk of serious complications of abdominal aortic aneurysm. Trials. 2021;22(1):962. doi:10.1186/s13063-021-05915-0

49. Pal R, Bhadada SK. AGEs accumulation with vascular complications, glycemic control and metabolic syndrome: A narrative review. Bone. 2023;176:116884. doi:10.1016/j.bone.2023.116884

50. Dhital S, Vyavahare NR. Nanoparticle-based targeted delivery of pentagalloyl glucose reverses elastase-induced abdominal aortic aneurysm and restores aorta to the healthy state in mice. Bader M, ed. PLOS ONE. 2020;15(3):e0227165. doi:10.1371/journal.pone.0227165

51. Schack AS, Stubbe J, Steffensen LB, Mahmoud H, Laursen MS, Lindholt JS. Intraluminal infusion of Penta-Galloyl Glucose reduces abdominal aortic aneurysm development in the elastase rat model. Bader M, ed. PLOS ONE. 2020;15(8):e0234409. doi:10.1371/journal.pone.0234409

52. Nectero Medical, Inc. Randomized Controlled Clinical Trial (RCT) of the Nectero EAST System for Small to Mid-Sized Abdominal Aortic Aneurysms (AAA) Stabilization: Evaluation of Efficacy. https://clinicaltrials.gov/study/NCT06001918

