## Supplementary Figure for "Distinct Type 2 Diabetes Components Exert Contrasting Effects on Abdominal Aortic Aneurysm Development"

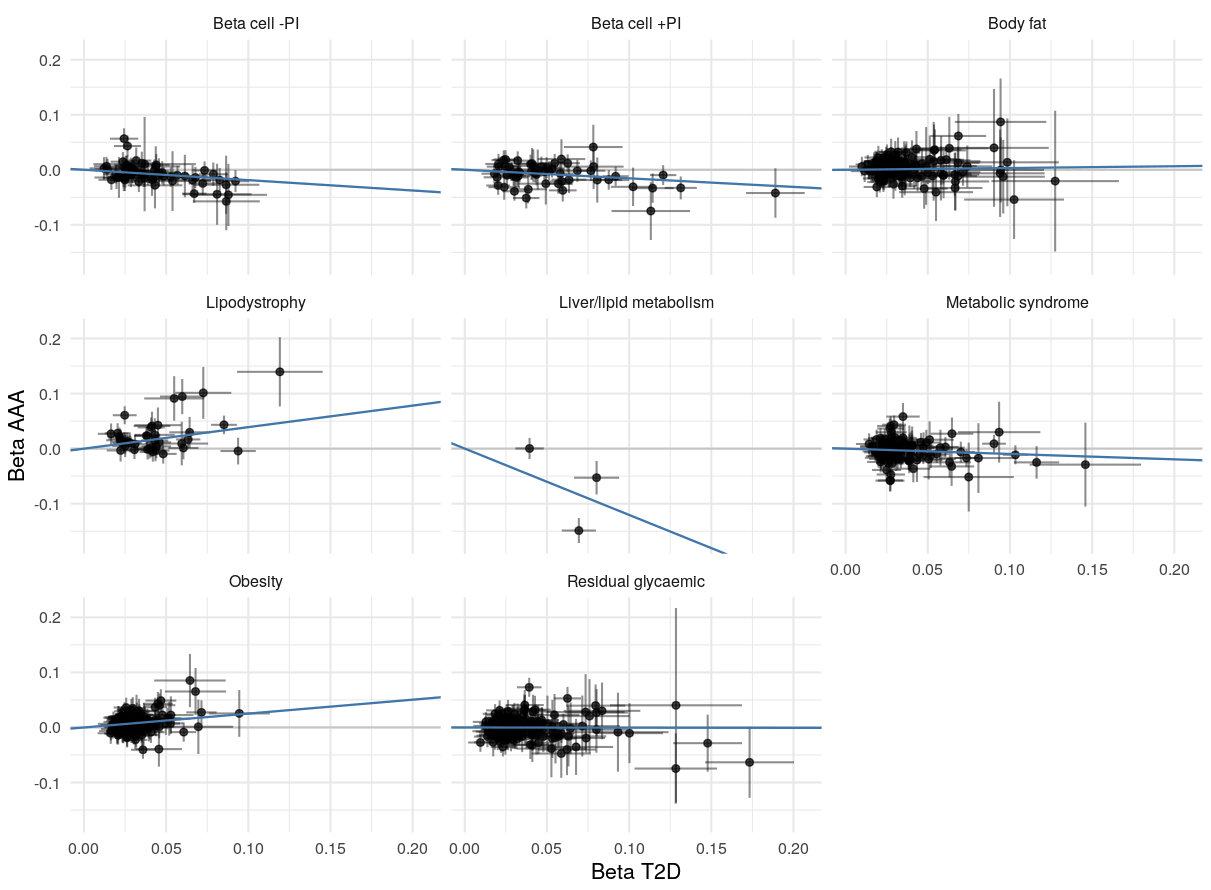

**Supplementary Figure 1. Plots of all genetic instruments per T2D cluster.** Scatter plots of AAA effect estimates against T2D effect estimates for variants within each cluster. The blue line denotes the cluster-specific IVW estimate of the effect of T2D on AAA.

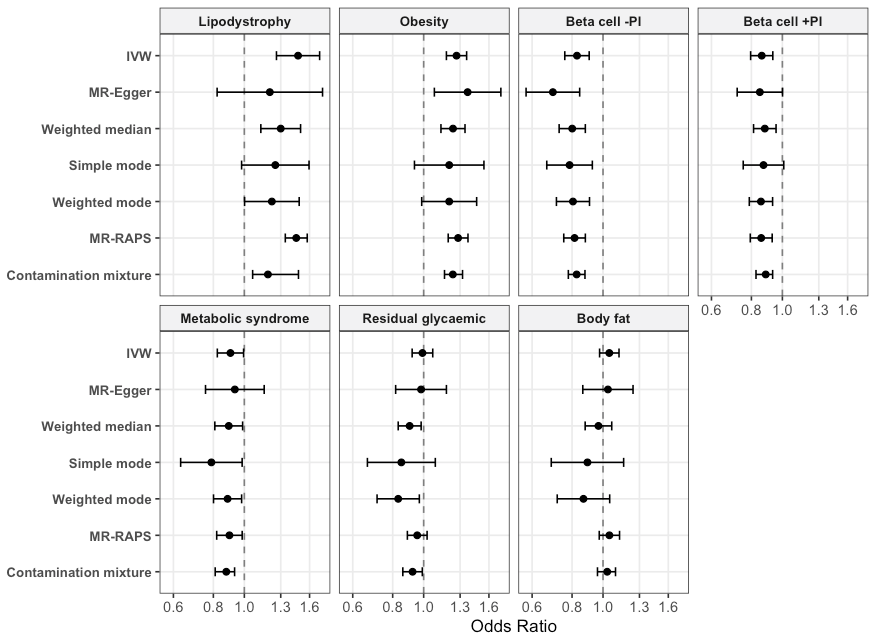

**Supplementary Figure 2**. **Effect estimates of Suzuki et al. T2D hard clusters on AAA across sensitivity analyses.** Effect estimates for the effect of T2D on AAA are reported for each cluster with 95% confidence intervals across sensitivity analyses, with odds ratios reported for AAA per unit log odds T2D. The Liver/lipid metabolism cluster only comprised 3 SNPs and is not included here.

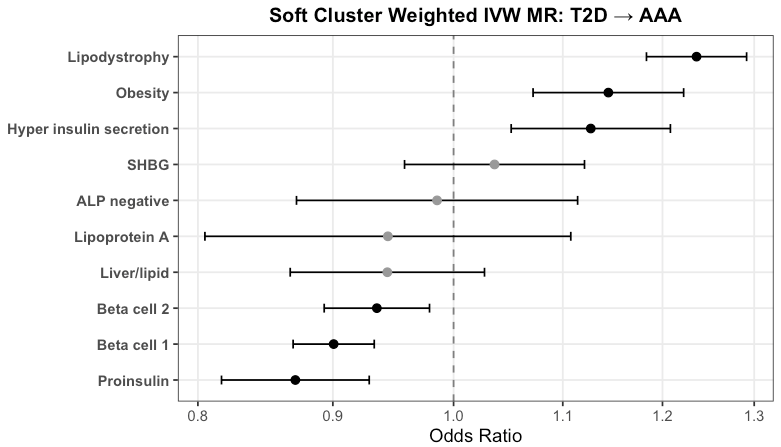

**Supplementary Figure 3. Effect estimates of Kim et al. T2D soft clusters on AAA.** The soft cluster weighted IVW effect estimates for the effect of T2D on AAA are reported for each cluster with 95% confidence intervals, with odds ratios reported for AAA per unit log odds T2D.

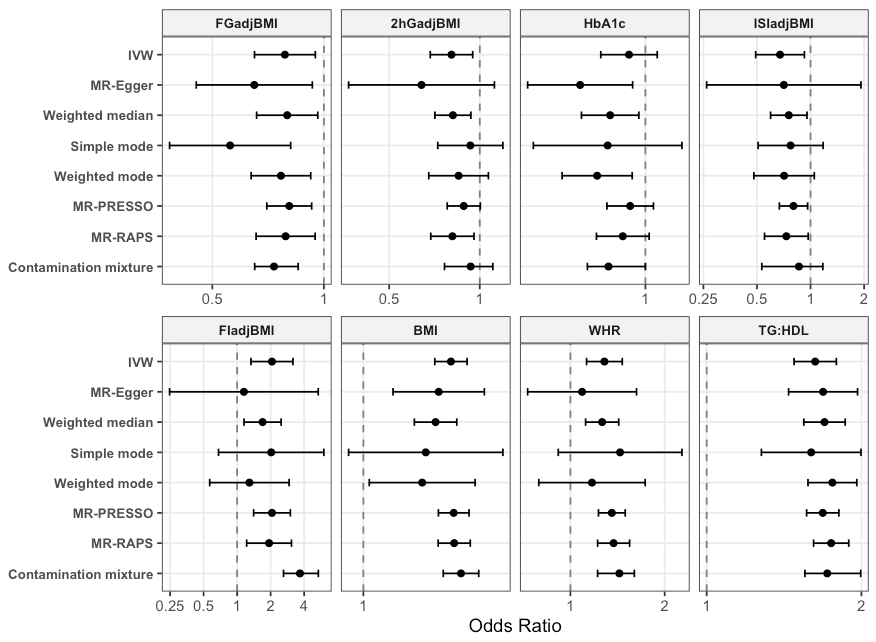

**Supplementary Figure 4. Effect estimates of glycemic traits on AAA across sensitivity analyses.** Effect estimates for glycemic traits on AAA are reported with 95% confidence intervals across sensitivity analyses. Traits are divided into those indexing hyperglycemia (Hemoglobin A1c, 2-hour glucose adjusted for BMI, and fasting glucose adjusted for BMI), insulin resistance (fasting insulin adjusted for BMI, triglyceride-to-HDL ratio, BMI, waist-to-hip ratio), and insulin sensitivity (modified Stumvoll insulin sensitivity index). Effect estimates for different glycemic traits are in different units. Magnitudes of the estimates should not be compared across traits. Rather, direction of the effect on AAA is emphasized in this analysis.
